# Metagenomic sequencing of composite airplane wastewater for surveillance of emerging viruses

**DOI:** 10.64898/2026.01.29.26343714

**Authors:** Michael R. McLaren, Olivia S. Hershey, Ari N. Machtinger, Daniel P. Rice, Alexandra M. Simas, Cindy R. Friedman, Dawn Gratalo, Casandra W. Philipson, William J. Bradshaw

**Affiliations:** SecureBio, Cambridge, MA, USA; Media Laboratory, Massachusetts Institute of Technology, Cambridge, MA, USA; Ginkgo Bioworks, Boston, MA, USA; Centers for Disease Control and Prevention, Atlanta, GA, USA

## Abstract

Robust early warning of emerging viruses requires sampling populations that drive spread coupled with assays capable of detecting new viral variants or species. Untargeted viral metagenomic sequencing can, in principle, detect any virus, including completely novel ones. Composite airplane wastewater enables monitoring long-distance travelers from central collection points; however, the performance of untargeted viral metagenomic sequencing in this sample type remains unknown. In municipal wastewater, abundant sewer-associated microbes and ribosomal RNA depress viral relative abundance, limiting metagenomic sensitivity. We compared untargeted viral metagenomic sequencing of composite airplane wastewater with time-matched municipal wastewater from the Greater Boston area. Human viruses and other human-associated taxa had consistently higher relative abundance in airplane samples than municipal samples, while most sewer-associated taxa had lower relative abundance. An increased relative abundance of human viruses lowers the sequencing depth required to detect emerging pathogens, suggesting that metagenomic sequencing of composite airplane wastewater is a cost-effective method for pathogen-agnostic surveillance.

**Importance:** Long-distance air travelers spread viral pathogens globally, making them an ideal sentinel population for pandemic surveillance systems. Testing composite airplane wastewater offers a practical, noninvasive approach to monitoring the traveler population. However, current surveillance systems rely on tests targeting specific known pathogens, missing novel threats. Untargeted metagenomic sequencing can detect viruses known or novel, but remains expensive to implement at scale; in municipal wastewater, sewer-derived microbes tend to overwhelm human viruses in sequencing data. We investigated whether a hypothesized reduced sewer microbial load in airplane wastewater would lower the sequencing effort required for viral detection. Understanding the performance of metagenomic sequencing in this context informs the design of cost-effective early-warning systems for emerging pandemics.

## Introduction

Long-distance air travelers are important vectors for the spread of emerging viral pathogens, making them a valuable sentinel population for pandemic early warning (Wegrzyn et al. (2023), Li et al. (2023), Bivins et al. (2024), St-Onge et al. (2025)). Wastewater from long-haul flights is a promising noninvasive sample stream for monitoring travelers; however, sampling individual planes is expensive and logistically challenging (Li et al. (2023), Bivins et al. (2024), NASEM (2024), St-Onge et al. (2025)). A potential solution is composite sampling at airport triturators—devices that process airplane wastewater before it enters the sewer system (NASEM (2024)). Triturators allow automatable collection of composite wastewater from large numbers of arriving flights with minimal operational disruption (NASEM (2024)). The use of multi-flight composite samples loses flight-level information (such as country of origin) but reduces processing costs and privacy concerns. Triturator monitoring for select respiratory pathogens is now routinely used at several airports as part of the US Centers for Disease Control and Prevention (CDC)’s Traveler-based Genomic Surveillance (TGS) program (Friedman, Morfino, and Ernst (2025), CDC (2025)).

Extant wastewater monitoring primarily uses pathogen-specific molecular assays, such as quantitative polymerase chain reaction (qPCR) and amplicon sequencing. However, broad-spectrum methodologies based on untargeted or semi-targeted metagenomic sequencing (MGS) can also be used to detect a wide range of enteric and non-enteric viruses in wastewater (Rushford et al. (2025), Tisza et al. (2023)). Semi-targeted MGS enriches for known viruses while tolerating some divergence, whereas untargeted MGS can theoretically detect entirely novel viruses. Such pathogen-agnostic capabilities are ideal for pandemic early warning.

However, deployment of wastewater MGS faces concerns about cost (NASEM (2024), Grimm et al. (2025)). Studies of municipal wastewater suggest the primary challenge is the low relative abundance of human viruses compared to the background microbial community; this high off-target background necessitates costly deep sequencing to detect viruses, especially non-enteric ones (Crits-Christoph et al. (2021), Rothman et al. (2021), Grimm et al. (2025), Rushford et al. (2025)). Probe-capture panels reduce the required depth by reducing this background, but only partially (Kantor and Jiang (2024)). A major background source is the sewer-system microbiome: Only 10–15% of microbial genetic material in treatment-plant influent is human-derived, with the remainder attributed to the sewer system itself (VandeWalle et al. (2012), Newton et al. (2015), McLellan and Roguet (2019), Fierer et al. (2022), Roguet et al. (2022)). Reducing this sewer component could substantially increase the relative abundance of human viruses.

Airplane wastewater, sampled closer to the point of excretion, likely contains fewer sewer-derived microbes than municipal wastewater. This reduction should lead to higher relative abundances of known and novel human viruses, reducing the sequencing depth needed for detection. However, the composition of airplane wastewater may be affected by other factors such as passenger lavatory habits (Jones et al. (2023)), low temperatures (Bivins et al. (2024)), elevated pH (Filho et al. (2017), Hjelmsø et al. (2019)), and the presence of lavatory treatment fluid (Bivins et al. (2024)). Only two studies have performed metagenomics on airplane wastewater (Nordahl Petersen et al. (2015), Hjelmsø et al. (2019)), neither using untargeted viral metagenomics. Further empirical study is needed to assess the performance of untargeted viral MGS of composite airplane wastewater and how it compares to the more widely studied municipal wastewater.

We applied untargeted viral MGS to composite airplane wastewater and compared it to time-matched municipal wastewater from the same city. Over a two-month period, we collected composite samples from an airport triturator and a nearby municipal treatment plant, processed samples with a viral-enrichment protocol, and measured samples using targeted assays and deep untargeted MGS of extracted RNA. Human viruses and other human-associated taxa had much higher relative abundance in airplane than municipal samples, while sewer-associated taxa were typically less abundant. These findings suggest that untargeted viral MGS would require substantially less sequencing effort to detect emerging human viruses in composite airplane wastewater than predicted from municipal wastewater studies.

## Results

### Sample collection and measurement

We collected samples between late October and December 2023 from the airport and treatment plant serving Greater Boston, Massachusetts, United States. Boston Logan International Airport (BOS) is the busiest airport in the New England region. Deer Island Wastewater Treatment Plant (DITP) is among the largest treatment plants worldwide, serving a population of approximately 2.3 million people across 43 municipalities (Duest (2023), U.S. Environmental Protection Agency (2023)). DITP’s catchment is split into North and South subsystems; BOS airport, which is located less than 3 miles away from the treatment plant, is served by the North subsystem. Airplane (BOS) samples represent the population of air travelers passing through or arriving to Boston, while DITP samples represent the local population of workers and residents. Parallel collection across the two sites enables direct comparison of traveler and local populations during the same period.

Routine collection of DITP samples began on October 31, 2023, with BOS sampling added on November 13, and continued through December 22. Sampling was performed on weekdays excluding holidays and unscheduled disruptions, resulting in 36 days of DITP data and 25 days of parallel BOS–DITP data (Figure 1, Table 4). BOS sampling was performed as part of the CDC’s TGS program. 24-hour composite samples of airplane wastewater were collected late morning. Each composite sample was collected via an autosampler from the inflow of a triturator which receives airplane wastewater from a large fraction of domestic and the majority of international arrivals. DITP sampling was performed mid-morning by treatment-plant staff, providing three distinct samples. Raw influent was collected separately from North and South sewer subsystems via autosampler as 24-hour composites. Primary sludge (settled solids) was collected from a combined (North and South) channel as a single grab sample. The two influent sources appear highly similar in our measurements; for simplicity, we group them together as “influent” unless otherwise specified. We refer to the three sample types as “airplane” (BOS triturator inflow), “sludge” (DITP combined sludge), and “influent” (DITP North and South influent).

**Table 1:** The estimated efficiency of ribodepletion at removing ribosomal versus non-ribosomal reads, by sample type. Estimates were obtained by linear regression on the log ratio of ribosomal to non-ribosomal reads, then exponentiating to convert back to the linear scale. Intervals are 95% parametric confidence intervals.

| Sample type | Estimate | 95% interval |
| --- | --- | --- |
| airplane | 120 | (380, 41) |
| influent | 20 | (28, 15) |
| sludge | 14 | (23, 8) |

**Table 2:** Conditions for RT-qPCR reactions.

| Step | Temp (C) | Time | Cycle |
| --- | --- | --- | --- |
| Reverse transcription | 50 | 5 min | 1x |
| RT deactivation | 95 | 20 sec | 1x |
| Denature | 95 | 3 sec | 40x |
| Anneal/Image (SARS-CoV-2) | 58 | 30 sec | — |
| Anneal/Image (PMMoV) | 60 | 30 sec | — |

**Table 3:** We prepared samples from four days in December for a pilot round of MGS.

| Collection date | Day of week | Handler |
| --- | --- | --- |
| 2023-12-05 | Tue | A |
| 2023-12-08 | Fri | B |
| 2023-12-12 | Tue | B |
| 2023-12-15 | Fri | A |

**Figure 1.**
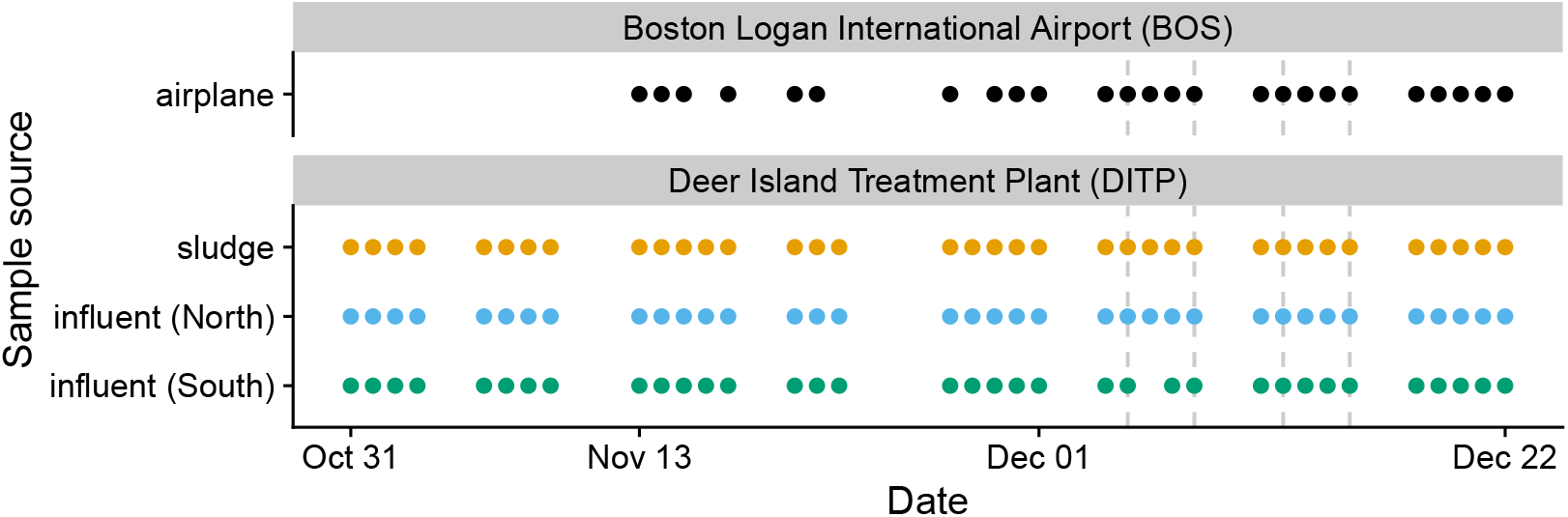
Sampling and processing schedule for a systematic comparison of metagenomic sequencing (MGS) of municipal and composite airplane wastewater. Samples were collected and processed on most weekdays from October 31 through December 22, 2023. Each collection day, samples were processed through total nucleic-acid extraction in one or two replicates. Each sample was assayed for total DNA and RNA concentrations and the concentrations of SARS-CoV-2 and pepper mild mottle virus; samples from four dates (dashed grey vertical lines) were additionally measured by untargeted viral MGS.

All samples were transported on ice and processed the same day through total nucleic-acid extraction using custom protocols designed to enrich for viruses over bacterial and human material. For each sample, we assayed total DNA and RNA concentrations and the concentrations of the viruses SARS-CoV-2 and pepper mild mottle virus (PMMoV). In addition, we performed a pilot round of untargeted viral MGS on samples from four dates in early-mid December (Figure 1, Table 3). To further enrich for viruses, sequencing preparation included a step to remove bacterial ribosomal RNA (ribodepletion); to assess the efficiency of this step, we sequenced each sample with and without depletion.

### Human viruses and other human-associated taxa have higher MGS relative abundance in airplane wastewater

For metagenomics to function as a cost-effective early warning system, pathogen RNA must be present at sufficient concentrations to be detected at practical sequencing depths. We therefore focused our analysis of MGS data on relative abundance, defined as the number of reads obtained from the target taxon per total reads sequenced. This metric serves as a proxy for detection efficiency: Any increase in a pathogen’s relative abundance yields a proportional reduction in the amount of sequencing required to detect it (Grimm et al. (2025)).

We compared viral relative abundances across sample types for 10 genera of human-infecting RNA viruses which were detectable in the four days of MGS data. These genera have diverse taxonomy and include viruses transmitted via respiratory and fecal-oral routes. Although highly variable, relative abundances across these diverse genera were consistently higher in airplane wastewater than in both municipal sample types (Figure 2). Comparing municipal sample types, abundances were generally similar between influent and sludge, though *Betacoronavirus* was higher in influent.

**Figure 2.**
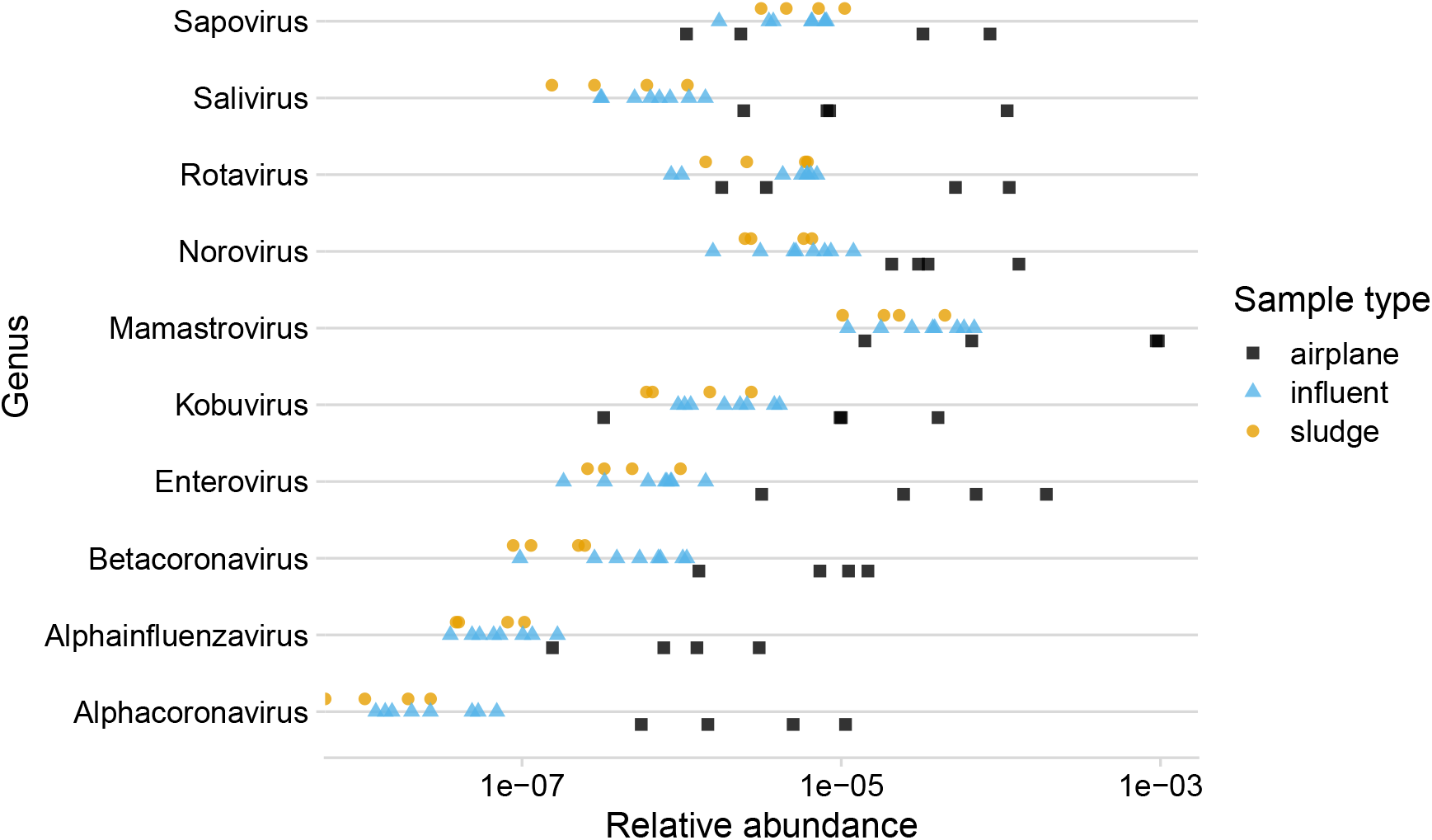
Across ten genera of human RNA viruses, viral relative abundances were typically greater in airplane than municipal wastewater. Points represent individual samples from the four-day metagenomic-sequencing pilot. The x-axis (log scale) shows relative abundance, defined as the ratio of deduplicated reads assigned to the taxon divided by total reads sequenced in ribodepleted libraries.

While higher prevalence in travelers might explain increases in relative abundance for specific viruses, the consistent elevation across 10 diverse genera suggests a systematic difference in sample composition. This observation supports our hypothesis that airplane wastewater contains significantly less material from the sewer microbiome, a reduction that would systematically inflate the relative abundance of human-derived taxa.

To further test this mechanism, we examined the relative abundances of additional human-associated taxa (dietary plant viruses, gut bacteria, and bacteriophages) and sewer-associated bacteria (Methods). For bacteria, we excluded ribosomal reads from the target counts (the numerator) to avoid confounding by variable ribodepletion efficiency, while retaining total sequenced reads as the denominator to maintain comparability to changes in human viruses. Relative abundances of human-associated taxa were universally higher in airplane samples (Figure 3). Conversely, 4 of 5 sewer-associated bacterial genera were lower in airplane samples, with only *Acinetobacter* showing similar abundance across sample types.

**Figure 3.**
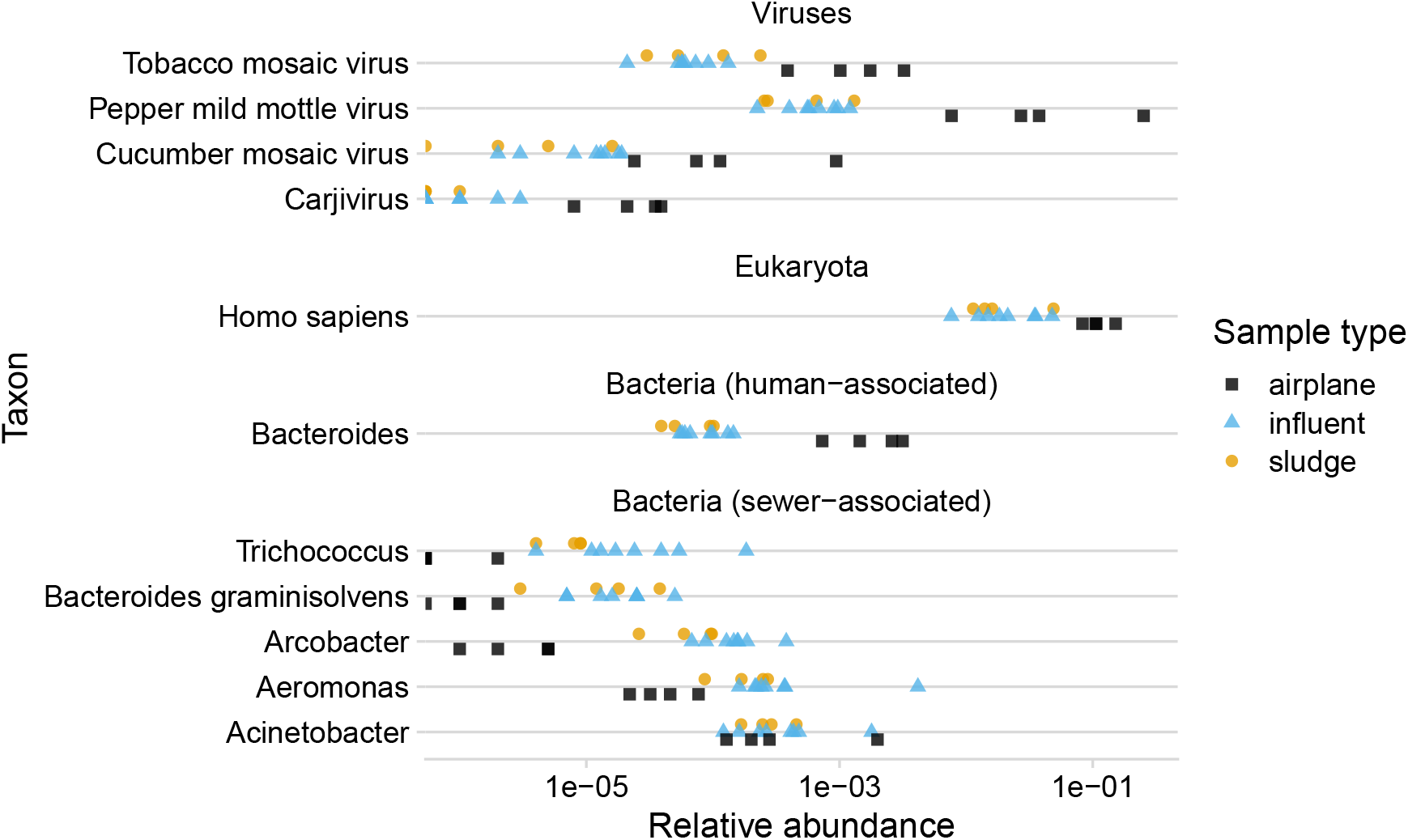
The relative abundances of non-pathogenic human-associated taxa are higher in airplane than municipal wastewater, while most sewer-associated bacteria are lower. Points represent individual samples from the four-day metagenomic-sequencing pilot. The x-axis (log scale) shows relative abundance, defined as the ratio of non-ribosomal reads assigned to the taxon divided by total reads sequenced. Ribosomal reads were excluded from the numerator to control for variable ribodepletion efficiency. Clipped points at the left edge indicate samples where there were zero assigned reads.

We quantified these differences using negative-binomial regression to estimate the fold-change in mean relative abundance between airplane wastewater and municipal influent (Figure 4, Table 6). Human viruses showed a 13-fold median increase in relative abundance in airplane wastewater (range: 5.5-fold for *Sapovirus* to 120-fold for *Alphacoronavirus*). Other human-associated taxa showed a similar enrichment, with a 24-fold median increase (range: 4.6-fold for *Homo sapiens* to 110-fold for PMMoV). In contrast, sewer-associated bacteria except *Acinetobacter* showed decreases of a similar magnitude.

**Figure 4.**
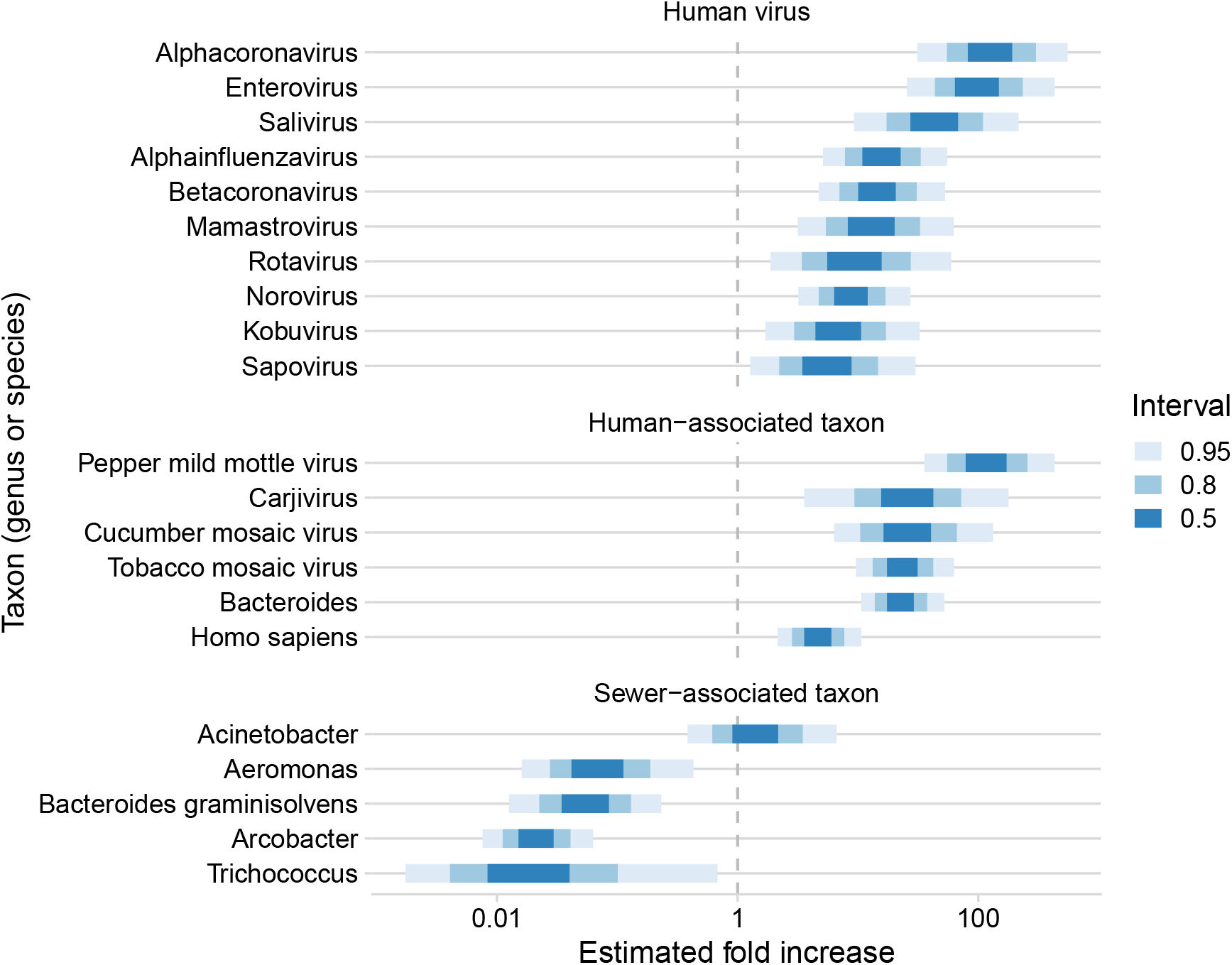
The fold increase in average relative abundance in airplane wastewater versus municipal influent is greater than one for human viruses and human-associated taxa and less than one for 4 of 5 sewer-associated taxa. Estimated fold increases are from Bayesian negative binomial regressions run individually for each taxon (4 airplane and 8 influent samples from 4 dates). The x-axis (log scale) shows posterior quantile intervals (50%, 80%, and 95% intervals).

These results suggest the presence of underlying differences in sample types that lead not just to increased relative abundance of human viruses, but also a wide range of human-derived taxa, and a similar scaled decrease for at least some sewer-derived microbes.

### High-level taxonomic differences between airplane and municipal wastewater

We next investigated whether differences in high-level taxonomic composition between sample types could further clarify the drivers of the increased relative abundance for particular human-associated taxa.

We first examined taxonomic composition at the domain level, distinguishing between ribosomal and non-ribosomal reads (Figure 5, Table 5). The domain-level composition of the three municipal sources look similar to each other and stand in marked contrast with airplane samples. While municipal samples are dominated by ribosomal reads, averaging 62% for sludge and 57% for influent, airplane samples average just 11%—more than 5-fold lower. For all other categories, airplane wastewater shows higher fractions. Compared to influent, airplane wastewater shows moderately higher fractions of non-ribosomal bacterial and unclassified reads (1.4-fold higher) and substantially higher fractions of eukaryotic (4.3-fold higher) and viral (8.1-fold higher) reads.

**Figure 5.**
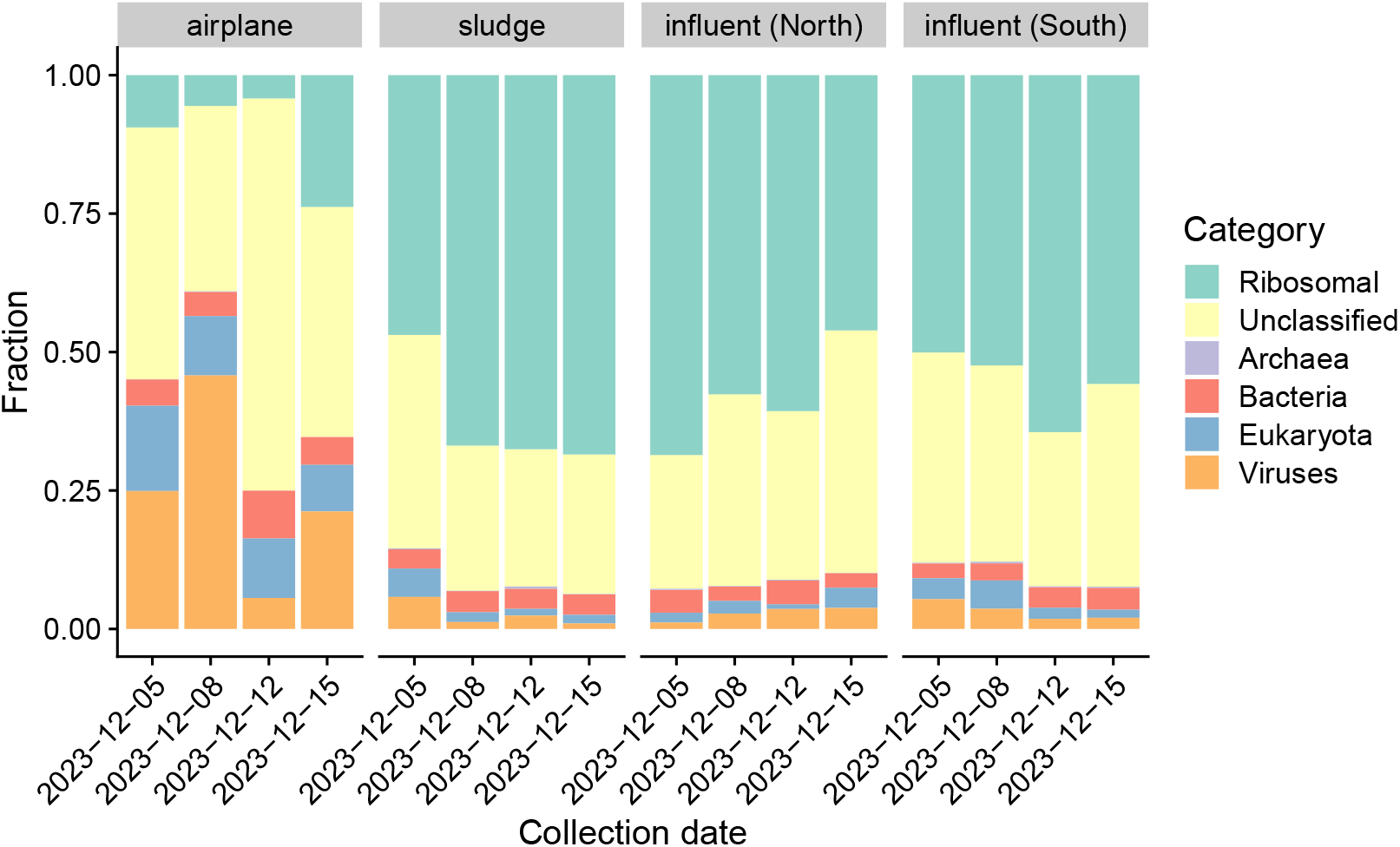
Airplane wastewater contains a significantly lower fraction of ribosomal reads and higher fractions of viral and eukaryotic reads compared to municipal wastewater. Domain-level taxonomic composition, estimated from a random subset of 1 million reads per sample using ribodepleted libraries, is shown for each sample grouped by sample source. The ‘Ribosomal’ category includes any reads identified as ribosomal regardless of taxonomic domain; all remaining categories only include non-ribosomal reads.

A decrease in ribosomal content could drive increases in other genomic features due to the sum-to-one constraint of fractional abundances (Gloor et al. (2017)). Supposing rRNA were to be removed from influent samples to bring the average fraction down to that in airplane samples, the average fractions of other categories would increase by a factor of 2.1. Therefore, variation in rRNA alone can potentially explain the full increase in non-ribosomal bacterial and unclassified reads, but only a small part of the increase in viruses overall or the increases in the relative abundances of human viruses and human-associated taxa seen in the previous section.

We next considered the composition of bacteria at the class level (non-ribosomal reads only; Figure 9) and viruses at the family level (Figure 10). Airplane and municipal wastewater show substantial overlap in the bacterial class composition, but with notable quantitative differences. In particular, airplane samples had larger fractions of *Clostridia* and *Actinomycetes* and smaller fractions of *Epsilonproteobacteria, Fusobacteriia*, and *Gammaproteobacteria* compared to municipal samples. These classes each contain ecologically diverse lineages, making detailed interpretation difficult; however, these differences suggest distinct compositions of bacteria between airplane and municipal wastewater.

Viral reads in all sample types are dominated by the family of plant viruses *Virgaviridae*; however, municipal samples also have large contributions from families in the class of RNA phages *Leviviricetes* which are rare in airplane samples. Airplane wastewater, on the other hand, shows greater fractions of the families *Astroviridae* and *Caliciviridae*, which contain human enteric viruses. These differences further support the idea that microbial RNA from airplane wastewater is primarily from direct human contributions, whereas that in municipal wastewater contains contributions from a distinct sewer environment.

### Bacterial ribosomal depletion was more effective in airplane samples

To clarify the cause of the lower levels of rRNA seen in airplane wastewater among ribodepleted samples, we compared the ratio of ribosomal to non-ribosomal reads in ribodepleted versus non-depleted samples. The ratio is moderately lower in airplane versus municipal wastewater in non-depleted libraries, but much lower in depleted libraries (Figure 6), indicating that the efficiency of ribodepletion at removing rRNA relative to non-rRNA was greater in airplane samples. We estimate ribodepletion efficiency to be approximately 6-fold greater in airplane versus influent samples (Table 1). Appendix 2 discusses possible causes for the lower baseline rRNA levels and improved ribodepletion efficiency seen in airplane wastewater.

**Figure 6.**
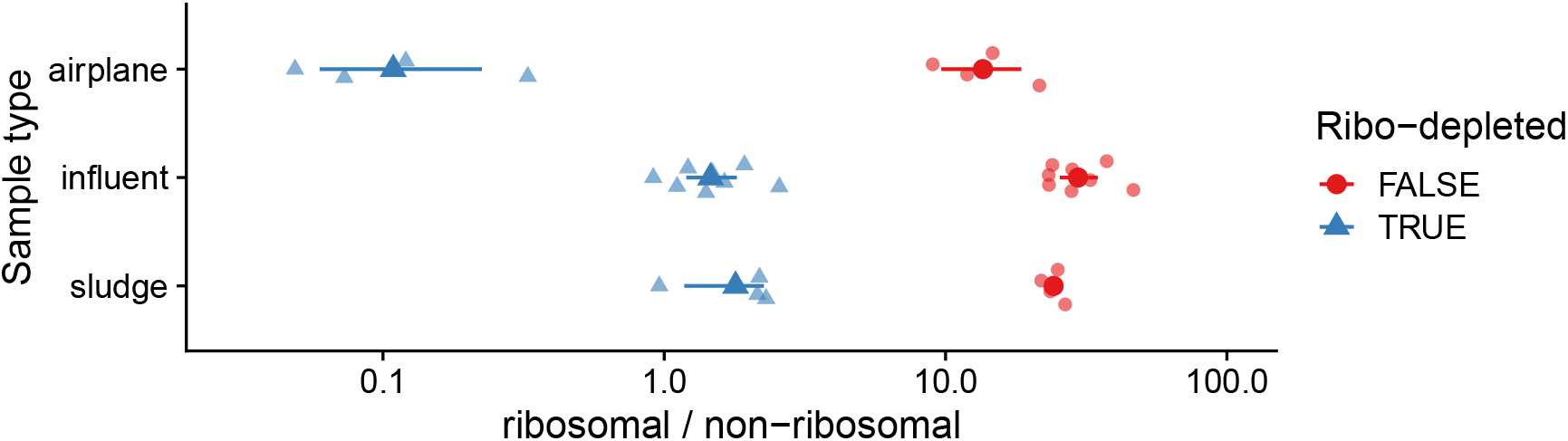
Airplane samples have lower baseline levels of rRNA and greater efficiency of ribodepletion than in municipal samples. Shown is the ratio of ribosomal to non-ribosomal reads (classified via k-mer matching to the SILVA database) for each sample (smaller semi-transparent points) along with their geometric means and 95% bootstrapped confidence intervals for each sample type and ribodepletion condition.

### Non-sequencing measurements suggest human-associated viruses form a higher fraction of total RNA in airplane wastewater

Our non-sequencing measurements provide an orthogonal view of relative abundances for SARS-CoV-2 and PMMoV over the full experimental time course. We estimated oncentrations of both RNA viruses via reverse transcription quantitative PCR (RT-qPCR) and the concentration of total RNA via Qubit florescence assay. We then computed the ratio of viral concentration to total RNA concentration for each virus. This ratio estimates the virus’ relative abundance as a fraction of total extracted RNA; variation in the ratio among samples is therefore predictive of variation in MGS relative abundances (Methods).

The ratio of viral-to-total RNA concentration for both SARS-CoV-2 and PMMoV is consistently higher in airplane wastewater than in both municipal sample types (Figure 7). For SARS-CoV-2, the ratio is also consistently higher in influent than in sludge. These observations are consistent with the patterns seen in MGS relative abundances for the 4-day subset (Figure 2, Figure 3).

**Figure 7.**
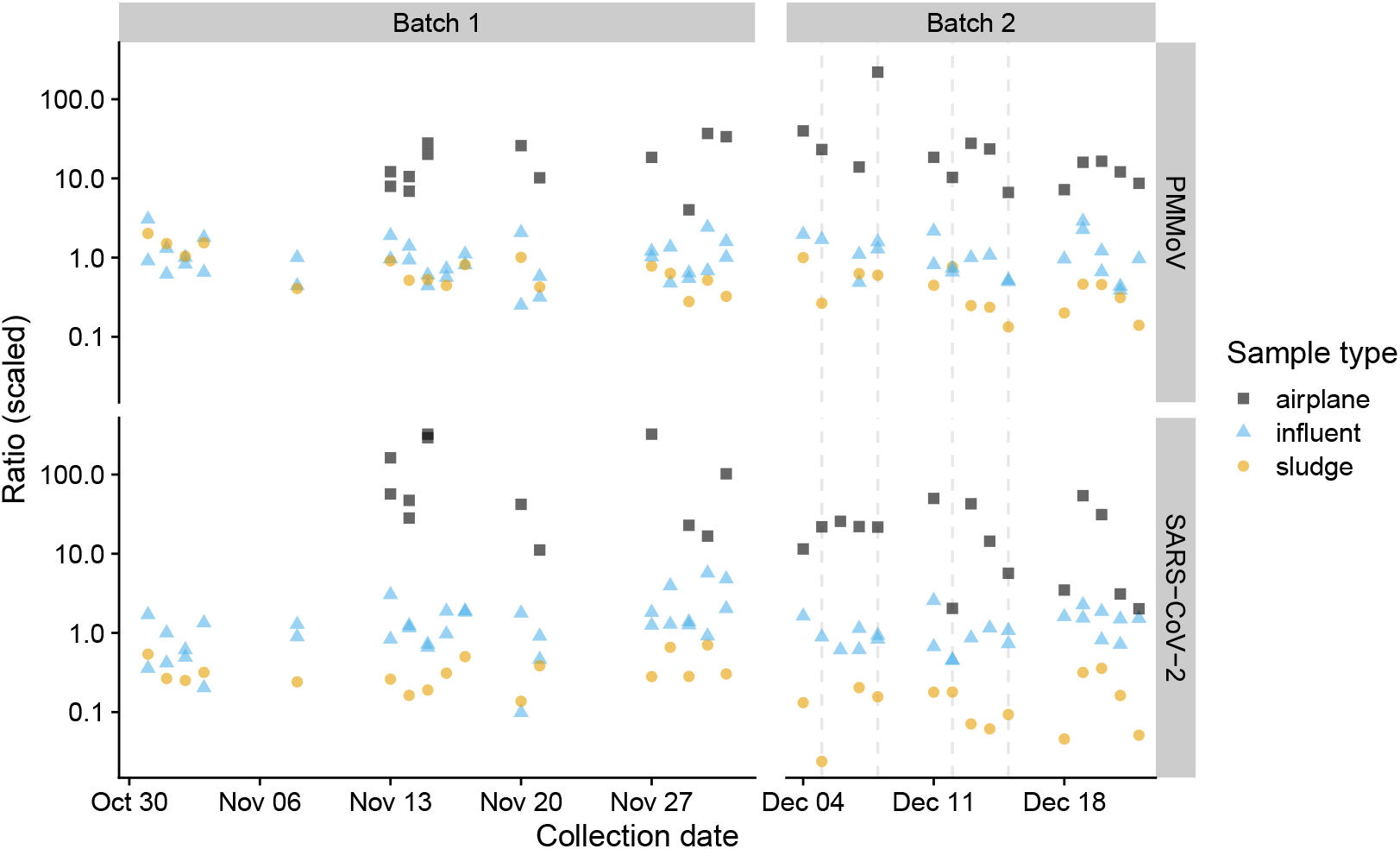
Estimated abundances of SARS-CoV-2 and PMMoV RNA relative to total RNA were consistently higher in airplane than municipal wastewater. The y-axis shows the estimated relative values among samples in the ratio of viral genome copies (measured by RT-qPCR) to total RNA (measured by Qubit). Qubit measurements for samples before and after December 4 are incomparable due to an experimental batch effect (Methods). Only the relative differences between samples for a given virus and Qubit batch are meaningful; ratios are scaled so that median value within a panel is 1. Vertical dashed lines indicate the four dates selected for metagenomic sequencing.

### Targeted measurements show distinct SARS-CoV-2 dynamics in airplane versus municipal wastewater

Our larger time-series of targeted measurements allows us to examine trends in SARS-CoV-2 infection dynamics which could impact how SARS-CoV-2 MGS relative abundance varies between sample types. To infer SARS-CoV-2 dynamics, we computed PMMoV-normalized SARS-CoV-2 concentrations from RT-qPCR measurements and compared them to positive test rates reported by the state of Massachusetts (Figure 8). Normalization to PMMoV accounts for variation in viral recovery and the amount of fecal input to the sample, improving the ability to infer trends from RT-qPCR measurements (Wolfe et al. (2021)). We note, however, that differences in normalized concentration between sample types are likely to reflect factors other than variation in SARS-CoV-2 infection rate. For example, normalized concentrations are consistently higher in influent than in sludge, consistent with previous findings that PMMoV partitions more strongly than SARS-CoV-2 to the solid versus liquid wastewater fraction (Kim et al. (2022), Barber et al. (2025)). Therefore our data do not allow us to infer differences in infection rates between traveler and local populations, only differences in their trends.

**Figure 8.**
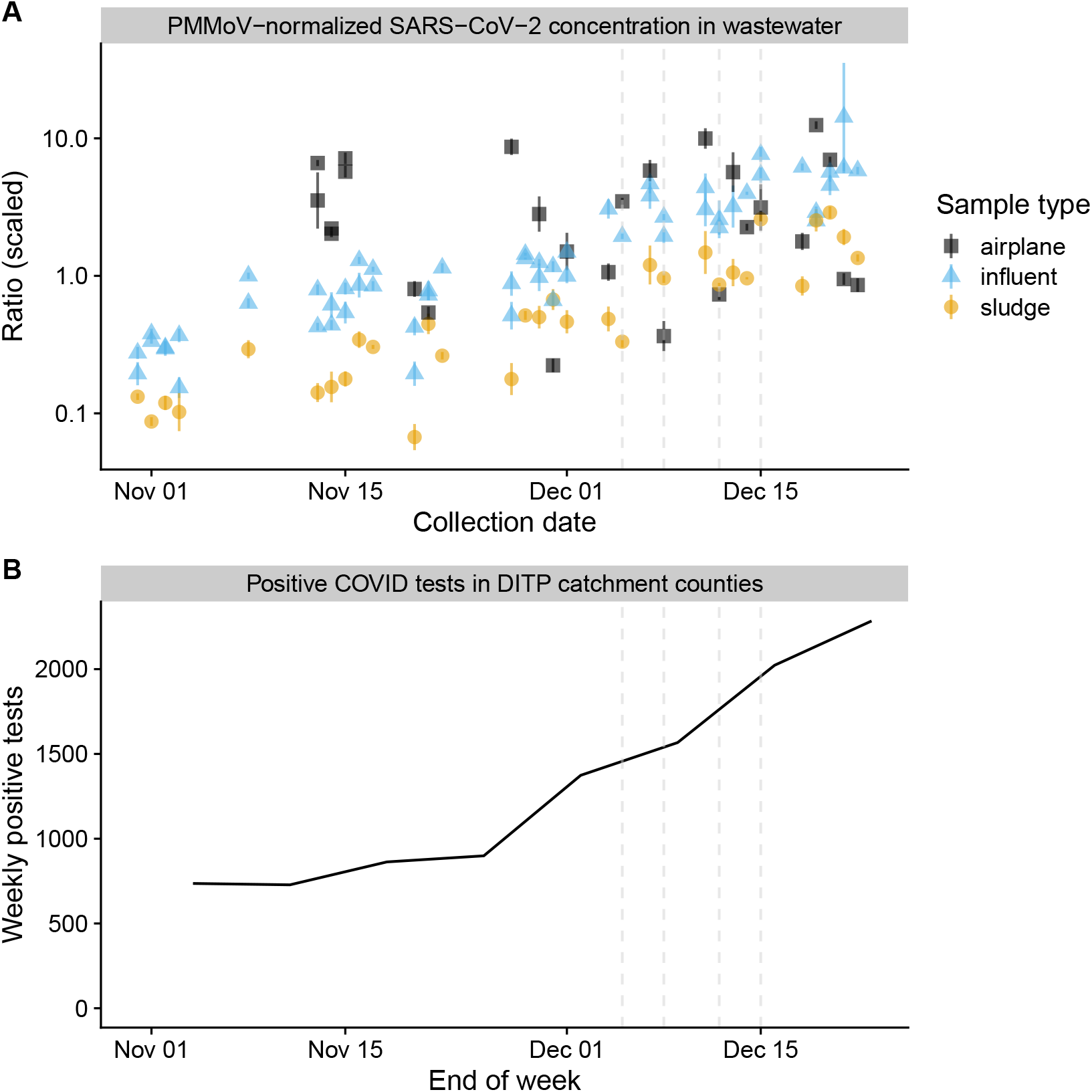
RT-qPCR measurements of SARS-CoV-2 normalized to PMMoV track reported COVID-19 infection trends for Greater Boston in municipal—but not airplane—wastewater. Panel A shows the ratio of SARS-CoV-2 to PMMoV (genome copies) estimated by RT-qPCR on collected wastewater samples. Only the relative differences between samples are meaningful; we therefore scale the ratios so that the median is 1. Panel B shows the number of positive COVID-19 tests from the past week reported by the Massachusetts Department of Public Health for the three counties (Suffolk, Norfolk, Middlesex) overlapping the treatment plant catchment. Vertical dashed lines indicate the four dates selected for metagenomic sequencing.

The PMMoV-normalized SARS-CoV-2 concentration in municipal influent and sludge steadily increased over the sampling period by over 10 fold (Panel A), while the weekly number of positive SARS-CoV-2 tests within the DITP catchment area increased by over 3 fold (Panel B). The normalized concentration seen in airplane wastewater (Panel A) is highly variable, but has a more stable average. These data suggests that the infection rate among travelers was roughly constant, but with high variance day-to-day, while the infection rate in the municipal population increased over the study period. Notably, normalized SARS-CoV-2 concentration in municipal, but not airplane, wastewater is higher on the pilot MGS dates than in early November. This observation suggests that the difference in *Betacoronavirus* relative abundances between airplane and municipal samples would have been greater for samples collected during this earlier period.

## Discussion

For the airport and treatment plant studied, relative abundance of human viruses in untargeted viral MGS data from composite airplane wastewater exceeded that in municipal wastewater by 5.5-to 120-fold (median 13-fold) across 10 diverse genera. Comparable increases occurred in non-ribosomal relative abundances for 7 other human-associated taxa, with decreases of a similar magnitude in 4 of 5 sewer-associated bacteria. While higher infection rates among travelers might explain increases in individual viruses, the consistent pattern across taxa suggests a systematic difference between sample types which elevates the relative abundance of human-associated taxa. We therefore predict that similar increases would apply for a novel, emerging virus.

These patterns can be largely explained by a lower concentrations of sewer-derived microbes. Previous estimates that 10-15% of microbial DNA in treatment-plant influent is human-derived (VandeWalle et al. (2012), Newton et al. (2015), McLellan and Roguet (2019), Fierer et al. (2022), Roguet et al. (2022)) suggest that eliminating the non-human component would increase relative abundances of human-derived taxa 6-to 10-fold. The effect could be larger for RNA, for example if metabolic activity is higher in sewer-than human-derived microbes.

The underlying mechanism may, however, be more complex. The reduced rRNA fraction in airplane samples was partly due to more efficient ribodepletion, potentially contributing a ∼2-fold increase in viral relative abundance. This efficiency difference could be explained by reduced RNA input in airplane samples or depletion probes having poorer coverage of sewer-than human-derived bacteria (Appendix 2). Before ribodepletion, airplane samples had much lower RNA concentrations (but not DNA) than municipal samples (Figure 12), suggesting selective RNA degradation. Since filtration removes intact cells, leaving primarily extracellular bacterial RNA, higher pH in airplane wastewater (Filho et al. (2017), Hjelmsø et al. (2019)) might selectively degrade cell-free bacterial RNA while preserving encapsulated viral RNA. Yet, this does not explain the 22-fold increase in the human-associated genus *Bacteroides*. Clarifying the mechanism could suggest future protocol improvements.

While human viruses consistently had higher relative abundance in airplane samples, the magnitude of the increase varied. Larger increases seen in some viruses (*Alphacoronavirus, Enterovirus*, and to a lesser degree *Salivirus*) could be explained by them having a greater difference between traveler and community infection rates. Our targeted SARS-CoV-2 measurements suggest that infection dynamics do in fact differ between these populations. However, we lack the necessary infection-rate information required to test this potential explanation. The heterogeneity we observed could alternatively be explained by differences is wastewater biochemistry (for example, how airplane lavatory treatment fluid interacts with viral particles) or bathroom behavior (for example, the relative mix of bodily fluids).

### Implications for the cost of MGS monitoring

Detecting a pathogen via MGS requires sequencing a sufficient number of pathogen reads across one or more samples. Following the model of Grimm et al. (2025), the expected number of pathogen reads for a given wastewater sample equals the total number of sequencing reads multiplied by the pathogen’s relative abundance. The relative abundance in turn is given by the pathogen’s rate of infection in the sampled population multiplied by a conversion factor specific to that pathogen, sample source, and MGS protocol. These conversion factors were previously measured by Grimm et al. (2025) for several viruses across studies which applied untargeted viral MGS to influent sampled from wastewater treatment plants. For example, they found that the average conversion factor across three studies for SARS-CoV-2, converting from weekly incidence (fraction of the population infected over the previous week) to relative abundance, was approximately 2 × 10^−6^. A pathogen whose genome sequence is known (such as SARS-CoV-2 by mid-January 2020) can be detected with as few as 3 reads by aligning reads to the genome (de Vries et al. (2021)). For a hypothetical weekly incidence of 10^−3^ (1 in 1000), this estimate implies that 1.5 billion total reads are typically needed to see 3 SARS-CoV-2 reads.

Our results lower expectations for the sequencing depth required for effective MGS early-warning systems using airplane wastewater. Without infection data, we cannot estimate conversion factors like Grimm et al. (2025). However, the higher relative abundances we observe across viruses provides strong evidence that conversion factors are typically higher in airplane than municipal samples for these sites and MGS protocols. Assuming the observed median 13-fold increase applies, the required sequencing for 3 SARS-CoV-2 reads drops from 1.5 billion to 120 million, significantly lowering sequencing costs, sample preparation needs, and computational burden. Importantly, larger conversion factors provide a cost and/or sensitivity benefit for MGS-based early warning that is independent of (and compounds with) any advantage in the form of higher infection rates among travelers which may be expected during the early stages of a pandemic.

Importantly, this analysis ignores the randomness inherent in pathogen detection. Airplane wastewater pools waste from fewer individuals than treatment plants, increasing the risk of missing pathogens if no infected individuals contribute to the sample. The higher day-to-day variance in airplane viral abundances we observed likely reflects these lower contributor numbers at least in part (with the changing daily flight mix also likely responsible). Ensuring sufficient aggregate passenger numbers across samples can mitigate increased sampling variation to fully leverage higher conversion factors.

### Limitations and next steps

This study was limited to one airport and one treatment plant over two months, with MGS on four dates. Multi-site, multi-season MGS studies are required to confirm generalizability. Future work should also assess how increased relative abundances translate to improvements in detection capability. Incorporating estimates of infection rates in the sampled traveler populations would enable model-based prediction of detection performance for hypothetical outbreak scenarios (as done in Grimm et al. (2025) for municipal MGS).

Protocol optimizations to reduce the ratio of off-target RNA may reduce the gap we observe between sample sources. Improvement in the removal of intact bacterial cells, the digestion of unencapsulated RNA, or ribodepletion efficiency could reduce bacterial RNA concentrations. Such improvements may especially improve viral relative abundances in municipal samples, where the concentration of bacterial RNA is greater. Alternatively (or in addition), pan-viral probe-capture panels can be used to enrich viruses with sufficient sequence similarity to known human viruses prior to sequencing (Tisza et al. (2023), Kantor and Jiang (2024), Mechikoff et al. (2025), Grimm et al. (2025)). Prove-based enrichment is imperfect and tends to have a multiplicative effect on viral relative abundances (Kantor and Jiang (2024)); thus, we expect airplane wastewater would retain its relative abundance advantage, but the magnitude and/or practical consequences of the difference may be reduced. Alternatively, municipal MGS monitoring could specifically target sites with naturally lower relative concentrations of sewer-derived microbes. Yet a third approach would be to look for municipal sampling locations that naturally have a higher ratio of human-to sewer-derived microbes.

Compared to airplane wastewater, municipal wastewater provides greater population coverage and a more direct measure of what is happening in the local community. Therefore, MGS of airplane wastewater is a complement, not a replacement, of existing municipal wastewater monitoring programs. Targeted and semi-targeted methods can help overcome the low relative abundance of pathogens in municipal wastewater (at the cost of being pathogen agnostic). Future work should explore how to most effectively combine these two approaches.

## Conclusion

Composite sampling of airplane wastewater enables detecting the spread of pathogens across state and national borders as well as tracking global trends from a small number of sampling points. Our results suggest that untargeted MGS of airplane waste for pathogen-agnostic viral detection is more cost effective than expected from prior studies of municipal wastewater. Future work should evaluate the cost required for optimal deployment of composite airplane MGS to provide early warning for emerging viral pandemics.

## Methods

### Sample collection

An overview of sample types and sampling locations and timeline are given in Results section “Sample collection and measurement”; here we provide further details.

#### Composite airplane wastewater from BOS airport

Composite airplane wastewater was collected from Boston Logan International Airport (BOS) as part of the US Centers for Disease Control and Prevention (CDC)’s Traveler-based Genomic Surveillance Program (Friedman, Morfino, and Ernst (2025)). An ISCO autosampler was positioned at the inlet of a triturator that receives waste from most international arrivals and a large share of domestic arrivals. Lavatory collection trucks routinely dispose of waste collected from one or more flights into the triturator to be broken down on its way into the sewer system. During each disposal event, the autosampler collected 50 mL every 2 min, collecting between 100-250 mL each time a truck was emptied, generating a 24-hour composite. During daily sample retrieval, the composite waste container was inverted to homogenize the contents, then poured through a funnel into collection bottles. Wastewater was collected by designated personnel at each location, transported and stored at 4 °C, and processed within 6 hours of collection.

#### Municipal wastewater from DITP

Deer Island Wastewater Treatment Plant (DITP) serves approximately 2.3 million people across 43 municipalities (Duest (2023), U.S. Environmental Protection Agency (2023)). The system feeding into DITP is a predominantly sanitary sewer system, with ∼5% being combined (waste- and stormwater) sewers. It is divided into North and South subsystems, with the North System receiving a larger daily flow (around 214 versus 114 million gallons per day). (BOS airport is served by the North system.)

#### We obtained three sample types

(i) 24-hour composites of raw influent from the North system, (ii) 24-hour composites of raw influent from the South system, and (iii) grab samples of primary sludge from a combined clarifier channel. North influent composites were typically pooled from East and West channel composites; on two dates, only one channel was available (East on 2023-11-27, West on 2023-12-21). South influent composites were collected from a single degritted channel; a South sample could not be collected on 2023-12-06. All municipal samples were prepared by treatment-plant staff in early-mid morning, delivered on wet ice the same morning, refrigerated on arrival, and processed within several hours of receipt.

### Sample processing

We processed samples with custom protocols designed with the joint aims of 1) enriching for viruses over bacterial and human cells and 2) treating each sample type similarly while also making type-specific adjustments to account for differences in concentration and consistency.

#### Particle removal and viral enrichment

Our protocols for each sample type involved treating samples with a detergent (Tween 20) to dissociate viruses from other particles, followed by a centrifugation (municipal samples only) and filtration to remove cells and other material larger than human viruses. For municipal influent samples, we treated 200 mL of influent by adding 10% Tween 20 to a final concentration of 0.1%, followed by vortexing at 1,000 rpm for 1 min, then sonicating at 40 kHz for 1 min. We then centrifuged the influent at 10,000g for 5 min to pellet large particles, and filtered the supernatant with a 0.45 m PES vacuum filter to remove remaining cells and larger solids.

Municipal sludge (DCS) samples were more concentrated than influent, being visibly less dilute and yield more total DNA and RNA (measured by Qubit as described below) than influent for a given amount of sample input volume. Therefore, we modified our influent protocol to use less input volume and a higher concentration of Tween 20. We diluted 20 mL of sludge with 20 mL phosphate-buffered saline (PBS) and increased the final concentration of Tween 20 to 0.5%, and then followed the vortexing, sonicating, centrifuging, and filtering protocol as described for influent samples.

Airplane wastewater was also more concentrated than influent; moreover, it had greater solid material (including paper debris) that led to clogging during vacuum filtration. For these samples, we diluted 40 mL of waste with 40 mL of PBS, brought the sample to a final concentration of Tween 20 to 0.5%, vortexed at full speed for 2 min, and sonicated as described for influent samples. In place of centrifuging, we added a 2 m glass microfiber pre-filter to the surface of the 0.45 m vacuum filter to remove large solids and other debris.

#### Concentration and extraction

For all sample types, we concentrated the viral-enrichment filtrate using the InnovaPrep Concentrating Pipette Select and revision E of the manufacturer recommended parameters for wastewater concentration using Ultrafiltration Concentrating Pipette Tips. We immediately added DNA/RNA Shield to the concentrated sample and incubated at room temperature for 30 min to prevent sample degradation by RNases and inactivate any viral particles. We extracted mixed nucleic acids from the total concentrated sample volume using the Zymo Quick-DNA/RNA Viral Kit, with reagent volumes scaled to maintain the reagent-to-sample ratios recommended by the manufacturer. Nucleic acids were eluted from the spin column using 50 L nuclease-free water and were divided into three aliquots and stored at -80C.

Airplane wastewater yields more DNA but less RNA per unit volume than both influent and sludge. See Figure 12 for DNA and RNA concentration measurements of samples processed with the final protocols.

#### Replicates and negative controls

We processed samples in duplicate when sample delivery time and lab schedule allowed (every processing date for airplane samples and roughly half of processing dates for municipal samples). Separate negative controls (PBS) were processed in parallel for each sample type during processing and viral enrichment, and were combined into a single daily negative control prior to nucleic acid extraction.

#### Sample handlers

Sample processing duties were split between two sample handlers (which we refer to as Handler A and Handler B), with a single handler responsible for receiving and processing the samples on any given day. Assignments were staggered across weekdays to avoid conflation of day-of-the-week and batch effects.

#### Detailed protocols

We provide detailed protocols on Protocols.io for each sample type: influent (Machtinger et al. (2025)), sludge (Machtinger, Hershey, Bradshaw, Soice, et al. (2024)), and composite airplane wastewater (Machtinger, Hershey, Bradshaw, Rice, et al. (2024)).

### Total and targeted nucleic-acid measurements

We performed total and targeted nucleic-acid measurements on at least one replicate for each sample type and date; these measurements provided quality control during the experiment and later served as independent measurements to compare to MGS data. Our total nucleic-acid measurements consisted of fluorescence-based (Qubit) assays of dsDNA and RNA concentrations. Our targeted measurements consisted of reverse transcription quantitative PCR (RT-qPCR) for SARS-CoV-2 and pepper mild mottle virus (PMMoV). The respiratory pathogen SARS-CoV-2 is readily quantifiable in wastewater via RT-qPCR (Duvallet et al. (2022)) and can be compared to local public health reporting. The plant virus PMMoV is heavily consumed by people through plant-derived food products and shed into wastewater, leading to frequent use as a marker of human fecal contamination and a quantitative measurement of human fecal contribution to wastewater samples (Symonds et al. (2018), Arts et al. (2023), Maal-Bared et al. (2023)).

#### Qubit

We determined the DNA and RNA concentrations for each sample type using the Qubit HS dsDNA assay and Qubit HS RNA assay, respectively. From 2023-10-31 to 2023-12-01, we performed Qubit assays using 2 L of undiluted sample from each replicate immediately after nucleic acid extraction. (We did not obtain dsDNA measurements on 2023-10-31.) After 2023-12-01, all Qubit measurements were performed weekly, using only one replicate per sample type per day. For these measurements, we made a 1:5 dilution of each nucleic acid sample, and used 2 L for each assay. These different procedures resulted in a strong batch effect in which later measurements gave higher estimated concentrations for all sample sources even after adjusting for volume differences (Figure 12). The remaining volume of the diluted samples was used for the RT-qPCR assays described below.

We estimated total DNA and RNA concentrations (in ng/ L) by adjusting the measurement from the Qubit instrument for the dilution used. Out-of-range measurements were set to the manufacturer’s reported detection limit for the given assay volume and likewise adjusted for dilution.

#### RT-qPCR

We used one-step RT-qPCR to measure the N2 region of SARS-CoV-2 using the CDC RUO primer and probe kit (REF, CDC 2020). The 20 L reaction contained the following components: 5 L TaqMan™ Fast Virus 1-Step Master Mix for qPCR (ThermoFisher cat: 4444434), 1.5 L of N2 assay in the SARS-CoV-2 Research Use Only qPCR Primer & Probe Kit (IDT, 10006713), 8.5 L RNase-free H2O, and 5 L of a 1:5 dilution of the extracted nucleic acid as the template RNA. We prepared a five-point standard curve using a quantitative SARS-CoV-2 RNA standard (VR-3276SD, ATCC), ranging from 1E0 to 1E4 copies per reaction. At least two no-template control reactions were run on each plate, and all sample and standard-curve reactions were prepared in triplicate.

We also measured PMMoV with RT-qPCR for use as an indicator of human fecal input and to demonstrate the effect on our protocol on viruses other than SARS-CoV-2. The 20 L one-step RT-qPCR reaction contained the following components: 5 L TaqMan™ Fast Virus 1-Step Master Mix for qPCR (ThermoFisher cat: 4444434), 0.3 L of the forward primer (GAG TGG TTT GAC CTT AAC GTT GA, 10 uM), 0.3 L of the reverse primer (TTG TCG GTT GCA ATG CAA GT, 10 uM), 0.2 uM of the probe (6-VIC-CCT ACC GAA GCA AAT G-MGB, 10 uM), 12.2 L of RNase-free H2O, and 2 L of a 1:5 dilution of the extracted nucleic acid as the template RNA. A standard curve was prepared with a quantitative PMMoV RNA standard (AM2070, Promega), ranging from 8E1 to 8E5 copies per reaction. At least two no-template control reactions were run on each plate, and all sample and standard curve reactions were prepared in triplicate.

For each virus and sample, we averaged the quantification cycle, or *C*_*q*_ value, across the three technical replicates. For PMMoV, one sample of each sample type was excluded for having only two valid replicates. We used standard curve reactions to qualitatively assess the validity of qPCR measurements, but did not perform a standard curve calibration. Instead, we estimate the viral concentration relative to a fixed, arbitrary baseline as 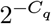, effectively assuming perfect qPCR efficiency. For this reason, our analysis is only semi-quantitative and only relative differences between samples for a given virus are interpretable.

#### Estimating relative abundances from RT-qPCR and Qubit measurements

To assess how the ratio of target viruses to total RNA varied among samples (Figure 7, we divide the value 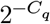 from RT-qPCR viral concentration estimates by the total RNA concentration (in ng/ L) measured by Qubit. Only relative differences between samples for a given virus and Qubit batch are interpretable. To facilitate visualization, we scale ratios by dividing by the median value for the given virus and Qubit batch.

Assuming that the ratio of the RT-qPCR target gene to all RNA from the virus is constant, then variation in this ratio among samples is expected to reflect variation in the fraction of total RNA that is derived from the given virus. Thus it is also expected to reflect variation in viral relative abundance we expect in our MGS data. However, we note some caveats. First, our RT-qPCR and Qubit measurements were performed on nucleic-acid extractions which were not subjected to the ribodepletion treatment performed during MGS preparation. Second, these measurements are not subject to the sequence deduplication performed for our SARS-CoV-2 MGS measurements. Third, RT-qPCR, Qubit, and MGS each have distinct sources of experimental bias that impact the resulting measurements. Multiplicative bias in measurements that is consistent across samples does not impact relative comparisons; however, such consistency is difficult to verify.

#### Estimating PMMoV-normalized SARS-CoV-2 concentrations

To estimate the abundance of SARS-CoV-2 relative to PMMoV (Figure 8), we first estimate Δ*C*_*q*_ (*C*_*q*_ of PMMoV minus *C*_*q*_ of SARS-CoV-2) for each sample via linear regression on RT-qPCR replicates across both viral targets. We then estimate the ratio of genome copies of SARS-CoV-2 to PMMoV by 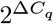. As noted above, this estimate assumes optimal qPCR efficiency and only provides the ratio compared to an arbitrary baseline. Only relative differences between samples are interpretable; for visualization, we scale ratios by dividing by the median value across all samples.

### Sequencing

For a pilot round of MGS, we selected four days in December across two weeks which were evenly split between two distinct days of the week (Tuesday and Friday) and our two different sample handlers (Table 3).

In prior analysis of comparable public wastewater datasets (Crits-Christoph et al. (2021), Rothman et al. (2021); analysis not shown) indicated that, even with steps taken to remove bacteria prior to extraction (including both filtration and bacterial ribodepletion), we observed that bacterial rRNA tends to dominate MGS data, with eukaryotic rRNA being a relatively minor component. We therefore chose to include a bacterial ribodepletion step during library preparation. To assess the efficacy of bacterial ribodepletion, we sequenced each sample with and without ribodepletion, resulting in 32 total sequencing conditions (4 dates × 4 sample sources × 2 ribodepletion states). Analysis of the composition of ribodepleted and non-depleted samples confirmed the dominance of bacteria among rRNA sequences and the utility of bacterial ribodepletion for reducing these sequences (Appendix 2).

Prior to submission for sequencing, we treated a single 15 L aliquot of purified total nucleic acids to remove DNA using the Invitrogen TURBO DNA-free Kit according to manufacturer’s instructions, then split the aliquot in two. We then submitted these 32 aliquots for library preparation (including ribodepletion) and sequencing to the MIT BioMicro Center. Aliquot’s receiving ribodepletion were treated with the [NEBNext rRNA Depletion Kit (Bacteria)](https://www.neb.com/en-us/products/e7850-nebnext-rrna-depletion-kit-bacteria). Libraries were then prepared for sequencing using the [NEBNext® Ultra™ II Directional RNA Library Prep Kit](https://www.neb.com/en-us/products/e7760-nebnext-ultra-ii-directional-rna-library-prep-kit-for-illumina) and sequenced on the Illumina NovaSeq 6000 platform, generating 2×150bp reads. An incident during library prep required several of the depletion preps to be repeated. The sequencing center redid all depletion preps, resulting in two depleted libraries for 9 of the 16 samples, which we combined the data from prior to our MGS data analysis. In total, these data comprised 3.3B read pairs (RNA; 2.1B with ribodepletion and 1.2B without).

Non-depleted libraries were dominated by rRNA, and are ill-suited for pathogen monitoring. Therefore we only consider the ribodepleted libraries in our main results. However, the overall efficacy of depletion and its differences between sample types are considered in Appendix 2.

### MGS data analysis

We analyzed data using the Nucleic Acid Observatory’s [Viral MGS Analysis Pipeline (v2.3.0)](https://github.com/naobservatory/mgs-workflow/tree/2.3.0), which proceeds as follows:

1. *Preprocessing:* Raw demultiplexed sequencing data (demultiplexed paired FASTQ files) are preprocessed through FASTP to remove adapter-contamination and low-quality sequences. Raw and FASTP-cleaned reads are run through FASTQC and MultiQC to check sequencing quality.
2. *Shallow taxonomic profiling:* Preprocessed read pairs from each sample are randomly subset to 1 million read pairs, which are then partitioned into ribosomal & non-ribosomal sequences with BBDuk using the SILVA SSU and LSU version 138.1 databases. Both ribosomal and non-ribosomal sequences then undergo taxonomic classification with Kraken2 (using the Standard reference database) to get an overall taxonomic breakdown for the sample.
3. *Deep human-virus profiling:* In parallel to (2), all preprocessed reads from each sample are screened for human-infecting virus reads as follows:
  1. Read pairs are aligned with Bowtie2 against a custom database of human-virus genomes generated as described in Grimm et al. (2025): “by obtaining all human-infecting virus taxonomy identifiers from Virus-Host DB (37); expanding this list to include all descendant identifiers; downloading all viral genomes corresponding to these identifiers from Genbank (38); and filtering the resulting database to remove transgenic and contaminated sequences.”
  2. Putative human-viral read pairs are then filtered by screening against known common contaminants (human, cow, pig, mouse and E. coli, as well as various genetic engineering vectors) using Bowtie2, discarding matching reads.
  3. Surviving read pairs are merged into single sequences, deduplicated, and run through Kraken2 using the Standard database. Reads that Kraken2 assigns to non-HV taxa are discarded.
  4. Finally, read pairs are annotated as human-viral if they either (a) are assigned to HV taxa by both Bowtie2 and Kraken2, or (b) are unassigned by Kraken2 and exceed a Bowtie2 alignment score threshold (alignmentScore/ln(readLength) >= 20).
  5. Each human-viral read pair is assigned an NCBI taxonomy ID (taxid) corresponding to the best alignment found by Bowtie2.
  6. The total number of read pairs assigned to a human-virus genus are found by summing the counts for all taxids descending from that genus’s taxid.

The taxonomic and human-virus profiling workflows each have limitations which affect our downstream use and interpretation of their results. The taxonomic profiling workflow uses the Kraken2 Standard database, which is targeted at prokaryotic and viral identification. It contains RefSeq genomes from archaea, bacteria, and viruses, along with plasmids, human, and UniVec Core (vector contamination) sequences included primarily to reduce prokaryotic and viral misclassification. Among eukaryotes, only the human (*Homo sapiens*) reference genome is included, along with some plasmids from various other eukaryotic organisms. Among non-ribosomal read pairs classified as *Eukaryota*, 99% are classified to *Homo sapiens*; however, due to the limited database we are unable to confidently say that read pairs classified to *Homo sapiens* are in fact derived from humans rather than other eukaryotes that could plausibly shed RNA into wastewater (for example, rats or other animals, or even fungi). Moreover, RefSeq is a relatively limited set of prokaryotic and viral genomes–read pairs from prokaryotes and viruses not represented in the database are likely to be misclassified to their closest relative or simply left unclassified.

The human-virus profiling workflow aims to overcome these database limitations by using an expanded set of human virus genomes from the much larger GenBank database. It also uses alignment-based mapping (with Bowtie2), which tends to be more sensitive and specific than Kraken2’s k-mer-based mapping. However, this combination is not without drawbacks. Our pipeline assigns read pairs to the taxid associated with the Bowtie2 best match (with random tie breaking), regardless of whether genomes from other taxids are similarly good matches. As a result, our pipeline may assign read pairs to lower taxonomic levels (e.g. species or subspecies) than justified by the data. To mitigate this issue, we only use the genus associated with each read-pair assignment in the present analysis. Another issue is that, by only including human viruses in our Bowtie2 database, our pipeline can be expected to assign read pairs from animal viruses to their closely-related human-viral relatives. We do not expect animal viruses to be present in airplane samples; however, they plausibly could be in municipal samples. If a significant fraction of read pairs from municipal samples classified as human viral were in fact from animals, then the increased sensitivity in airplane wastewater presented in the Results would be an underestimate. Thus, the possible misclassification of animal viruses as human does not affect our overall conclusion.

#### Calculating relative abundances

Relative abundances of human viral genera were computed from deep human-virus profiling by dividing the number of deduplicated reads assigned to the genus by the total number of reads sequenced (prior to any quality filtering or deduplication). This definition is meant to reflect the amount of nonredundant information available for detecting a pathogen for a given amount of sequencing effort.

Relative abundances of other taxa were computed from shallow taxonomic profiling by dividing the number of non-ribosomal reads assigned to that taxon (without deduplication) by the number of reads in the subset that underwent shallow profiling. Here we avoid deduplication because 1) deduplication rates from a small subset are not representative of the full set of reads and 2) our aim with these taxa is to understand the taxonomic composition of the reads regardless of whether reads carry redundant genomic information.

### Selection of human- and sewer-associated comparison taxa

To evaluate our hypothesis that a reduction in sewer-associated microbes were driving variation in human virusus, we selected a range of comparison taxa that are associated with the human fecal microbiome or the sewer microbiome. The human-associated taxa include diet-derived plant viruses, prominent members of the human gut microbiome, and humans themselves (*Homo sapiens*). We included plant viruses from the genus *Tobamovirus* and several of its species: Pepper mild mottle virus (PMMoV), Tobacco mosaic virus, and Cucumber mosaic virus. From the gut microbiome, we include the bacterial genus *Bacteroides*, a dominant member of the human gut microbiome which is far less prominent in municipal wastewater. We also include the genus of DNA viruses *Carjivirus*, viruses that infect *Bacteroides* and are highly abundant in the human gut (Smith et al. (2023), Ramos-Barbero et al. (2024)). For sewer-associated taxa, we included four bacterial genera—*Acinetobacter, Aeromonas, Arcobacter*, and *Trichococcus*—found to be highly abundant in US and global studies of sewage communities and thought to reside in sewer pipes (McLellan and Roguet (2019)). We also include the species *Bacteroides graminisolvens*, which is related to prominent gut *Bacteroides* but has been found to be specifically associated with sewer pipes (Feng and McLellan (2019)).

### Public health comparison data

We obtained Massachusetts COVID-19 surveillance data from the Massachusetts Department of Public Health (MDPH) 2023–2024 archived COVID-19 reporting dashboard data (https://www.mass.gov/lists/archive-of-covid-19-data-in-massachusetts; downloaded 2025-05-25). We extracted county-level weekly positive-tests totals and aggregated Suffolk, Norfolk, and Middlesex counties to approximate the Deer Island Wastewater Treatment Plant catchment.

### Statistical data analysis and visualization software

Analysis and visualization of total (Qubit) and targeted (RT-qPCR) nucleic-acid measurements and of MGS pipeline results were performed in R (R Core Team (2025)) with tidyverse (Wickham et al. (2019)). Standard linear regression was performed with the lm function from base R. Bayesian regression modeling was performed with Stan (Team (2023)) via the rstanarm R package (Goodrich et al. (2024)) using default priors.

## Data and code availability

*Placeholder for statement of data and code availability. Code along with metadata, RT-qPCR measurements, and Qubit measurements will be published on GitHub. Raw MGS data with human reads removed will be published in the NCBI Sequence Read Archive (SRA)*.

## Appendix 1: Taxonomic composition

We first looked at high-level composition in terms of the fraction of reads classified as ribosomal and at the domain level (for non-ribosomal reads), as reported in the Results (Figure 5, Table 5).

We next looked to see if high-level differences could be observed in bacterial and viral composition. Substantial differences can be seen in class-level composition among non-ribosomal bacterial reads (Figure 9) between airplane and municipal wastewater. In particular, airplane samples show increased fractions of *Clostridia* and *Actinomycetes* and reduced fractions of other classes including *Epsilonproteobacteria, Fusobacteriia*, and *Gammaproteobacteria*.

**Figure 9.**
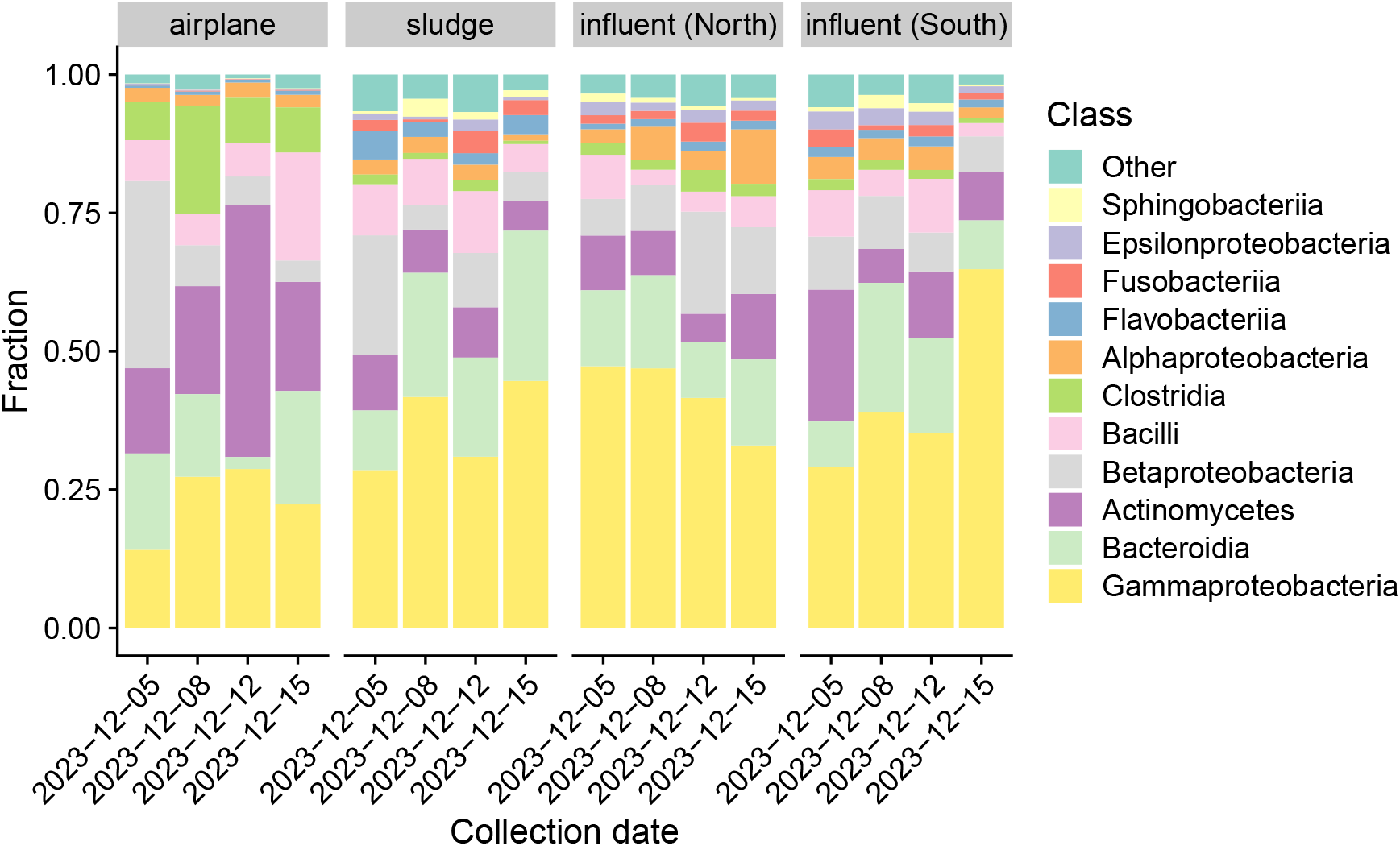
Bacterial class composition (non-ribosomal reads only) in ribodepleted libraries. Classes that are less than 0.02 in every sample are grouped as “Other.” Classes are ordered by average fraction across all samples.

The composition of viral families (Figure 10) among samples dominated by the family of plant viruses *Virgaviridae*; however, municipal samples also have large fractions from families of the *Leviviricetes* class of RNA phages, including *Fiersviridae, Steitzviridae, Blumeviridae*, and *Solspiviridae*. These RNA phages appear far less abundant in airplane samples. Excluding *Virgaviridae* (Panel B), we see that two families associated with human enteric disease, *Astroviridae* and *Caliciviridae*, appear much more prominently in airport than in municipal samples. *Alphaflexiviridae*, a family which infects plants and fungi, is apparent across all sample types but especially in one airplane sample.

**Figure 10.**
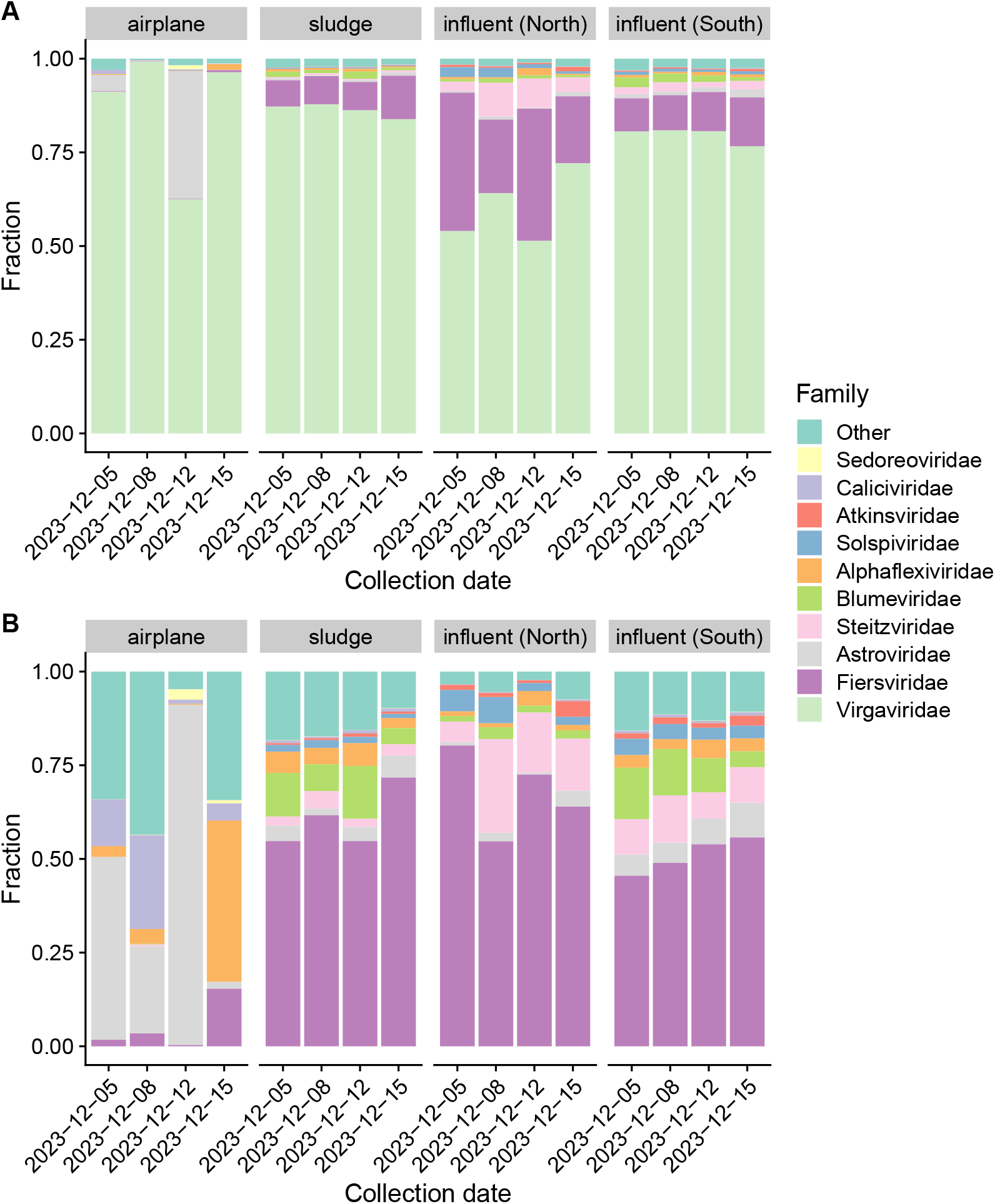
Viral family composition with (Panel A) and without (Panel B) the family of plant viruses *Virgaviridae*. Panel A shows fractions among all reads assigned to a viral family; families that are less than 0.01 in every sample are grouped as “Other.” Panel B shows the same families from Panel A, but with fractions recomputed after removing *Virgaviridae*. Families are ordered by average fraction across all samples, with most abundant (*Virgaviridae* in Panel A, *Fiersviridae* in Panel B) at the bottom.

## Appendix 2: Ribosomal composition and depletion

Taxonomic characterization of reads classified as ribosomal indicates that the vast majority of ribosomal RNA (rRNA) in our extractions is bacterial in origin. In non-depleted samples, an overwhelming majority of ribosomal reads are classified as bacterial (Figure 11, top row). In ribodepleted samples, the majority of reads are also classified as bacterial; however, we also see significant minorities classified as archaeal and eukaryotic and a large minority (nearly 50% in one sample) are left unclassified (Figure 11, bottom row), reflecting limits in the ability of our bioinformatics workflow to classify ribosomal reads.

**Figure 11.**
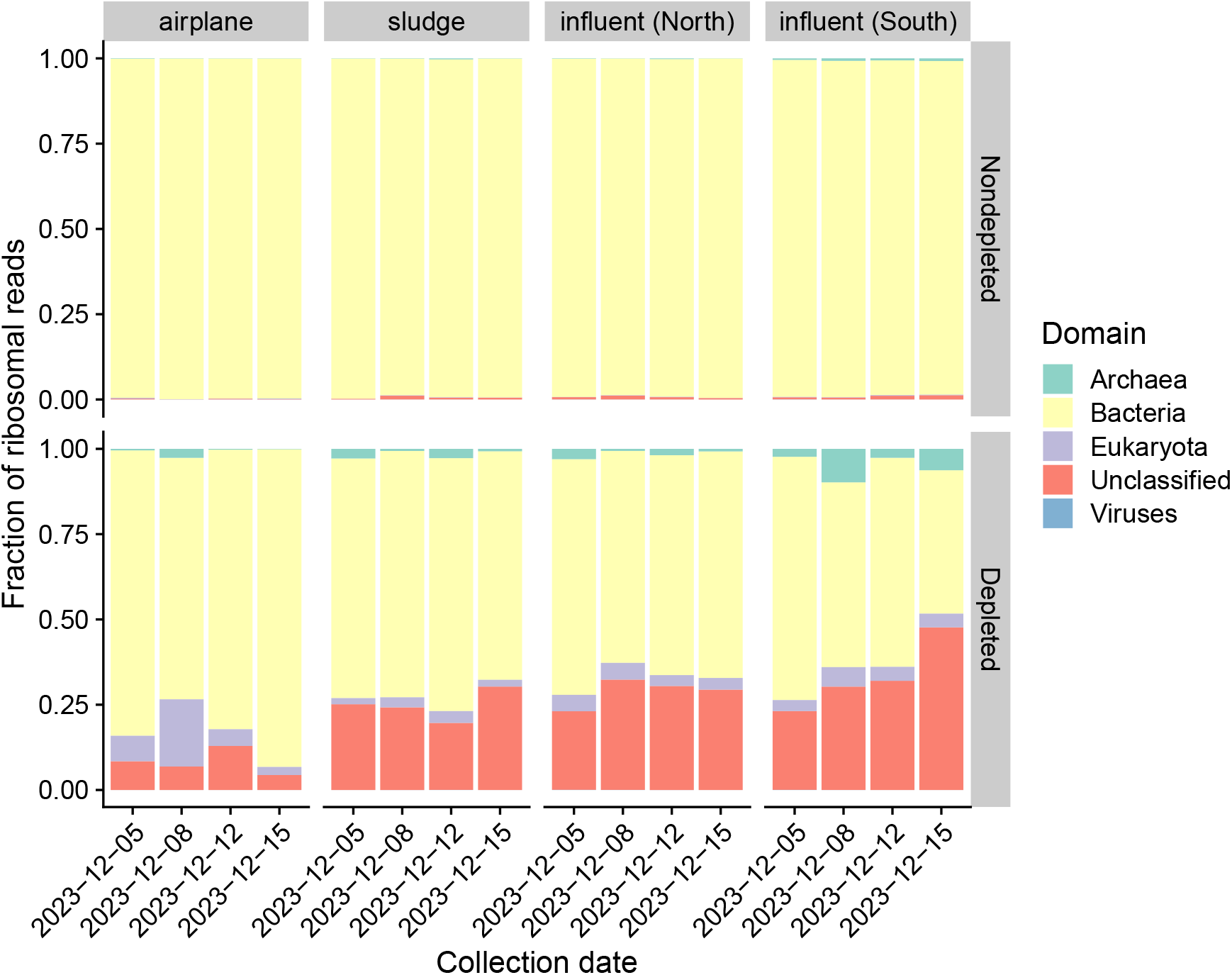
Domain-level taxonomic composition of ribosomal reads in ribodepleted and non-depleted libraries. Reads were classified as ribosomal by BBDuk based on k-mer similarity to sequences in the SILVA SSU and LSU database; then ribosomal and non-ribosomal reads were classified to the domain level with Kraken2. This figure shows the composition among just ribosomal reads for depleted and non-depleted libraries, whereas Figure 5 shows the composition among all (ribosomal and non-ribosomal) reads for just depleted libraries.

The lower baseline level of rRNA may be due to the hypothesized greater ratio of human to sewer microbial content in airplane samples; however, it is unclear whether the sewer microbiome intrinsically has a greater ribosomal fraction than the human microbiome. Another possible explanation lies in the interaction between our processing protocol and the distinct chemical environment of the airplane samples. As our protocol employs filtration to remove intact cells prior to extraction, most ribosomal reads may come from cell-free rRNA. The presence of airplane lavatory treatment fluid might cause this unprotected rRNA to preferentially degrade in airplane samples. Yet a third possibility is that the pre-filter used in our airplane protocol improves the removal of intact bacteria.

The greater ribodepletion efficiency (shown in Results) may also have multiple explanations. First, the lower baseline concentration of rRNA in airplane samples may lead to improved depletion performance. The combination of a lower fraction of rRNA out of all RNA, as seen in non-depleted samples in Figure 6, with a lower concentration of total RNA, as seen in Figure 12, imply that airplane samples have a much lower concentration of rRNA than municipal samples. Ribodepletion kits can become saturated at overly high rRNA levels, leading to reduced depletion performance—plausibly such saturation is occurring in municipal but not airplane samples. Second, the ribodepletion kit may have better coverage for bacteria in the airplane samples rather than in municipal samples. NEB provides limited information about the bacterial reference sequences used for the design of the probes in their kit. However, it is plausible that the references are biased towards bacteria found in the human rather than sewer microbiome, in which case we can expect ribodepletion to work less efficiently in the sewer-dominated municipal samples.

## Additional tables

**Table 4:**
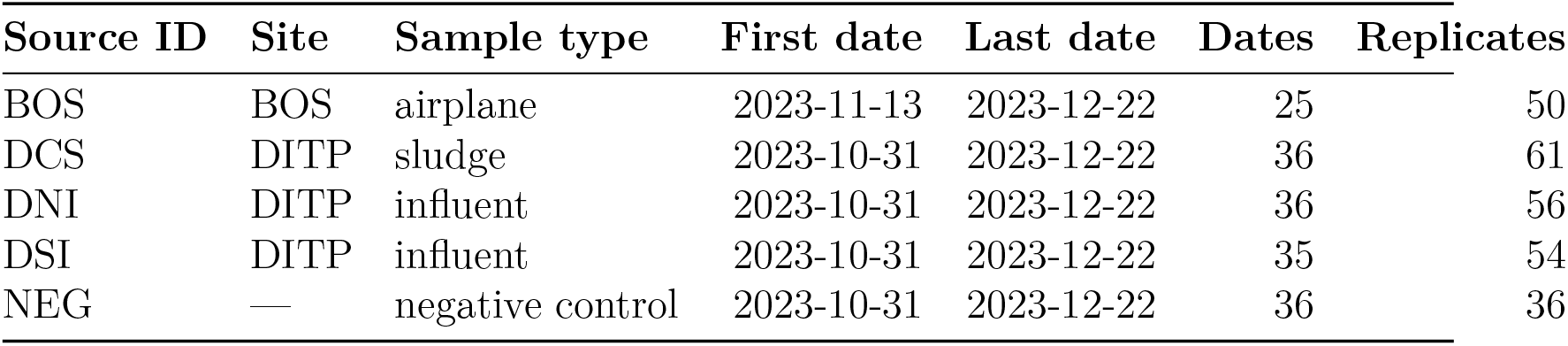
Samples were collected and processed in replicate from late October through late December 2023.

| Source ID | Site | Sample type | First date | Last date | Dates | Replicates |
| --- | --- | --- | --- | --- | --- | --- |
| BOS | BOS | airplane | 2023-11-13 | 2023-12-22 | 25 | 50 |
| DCS | DITP | sludge | 2023-10-31 | 2023-12-22 | 36 | 61 |
| DNI | DITP | influent | 2023-10-31 | 2023-12-22 | 36 | 56 |
| DSI | DITP | influent | 2023-10-31 | 2023-12-22 | 35 | 54 |
| NEG | — | negative control | 2023-10-31 | 2023-12-22 | 36 | 36 |

**Table 5:** Domain-level taxonomic composition estimated from a subset of 1M read pairs, as shown in Figure 5. Table shows the average fractions across samples from each sample source. The ‘Ribosomal’ category includes any reads identified as ribosomal regardless of taxonomic domain. All remaining categories only include non-ribosomal reads.

| Category | airplane | sludge | influent (South) | influent (North) |
| --- | --- | --- | --- | --- |
| Ribosomal | 0.11 | 0.62 | 0.56 | 0.58 |
| Unclassified | 0.48 | 0.29 | 0.34 | 0.33 |
| Archaea | 0.00036 | 0.0017 | 0.0020 | 0.0012 |
| Bacteria | 0.057 | 0.037 | 0.034 | 0.034 |
| Eukaryota | 0.11 | 0.024 | 0.031 | 0.021 |
| Viruses | 0.24 | 0.026 | 0.032 | 0.028 |

**Table 6:** Estimated fold increases in relative abundance in airplane versus municipal influent for various human viruses and reference taxa. Estimation was performed with a Bayesian negative binomial regression using the rstanarm package. The table shows the median and 50% and 95% quantile intervals.

| **Taxon** | **Estimate** | **50% interval** | **95% interval** |
| --- | --- | --- | --- |
| **Human virus** |  |  |  |
| Alphacoronavirus | 120 | (82, 190) | (31, 550) |
| Enterovirus | 97 | (64, 150) | (26, 430) |
| Salivirus | 43 | (27, 68) | (9.3, 220) |
| Alphainfluenzavirus | 15 | (11, 23) | (5.1, 55) |
| Betacoronavirus | 14 | (10, 21) | (4.7, 53) |
| Mamastrovirus | 13 | (8.2, 20) | (3.2, 62) |
| Rotavirus | 9.2 | (5.6, 16) | (1.9, 60) |
| Norovirus | 8.7 | (6.3, 12) | (3.2, 27) |
| Kobuvirus | 6.8 | (4.4, 11) | (1.7, 33) |
| Sapovirus | 5.5 | (3.4, 8.8) | (1.3, 30) |
| **Human-associated taxon** |  |  |  |
| Pepper mild mottle virus | 110 | (79, 170) | (36, 430) |
| Carjivirus | 26 | (16, 42) | (3.6, 180) |
| Cucumber mosaic virus | 25 | (16, 40) | (6.3, 130) |
| Tobacco mosaic virus | 23 | (17, 31) | (9.6, 63) |
| Bacteroides | 22 | (17, 29) | (11, 52) |
| Homo sapiens | 4.6 | (3.6, 6) | (2.2, 11) |
| **Sewer-associated taxon** |  |  |  |
| Acinetobacter | 1.4 | (0.9, 2.2) | (0.38, 6.6) |
| Aeromonas | 0.066 | (0.042, 0.11) | (0.016, 0.43) |
| Bacteroides graminisolvens | 0.055 | (0.034, 0.085) | (0.013, 0.23) |
| Arcobacter | 0.021 | (0.015, 0.03) | (0.0076, 0.063) |
| Trichococcus | 0.018 | (0.0083, 0.04) | (0.0017, 0.68) |

## Additional figures

**Figure 12.**
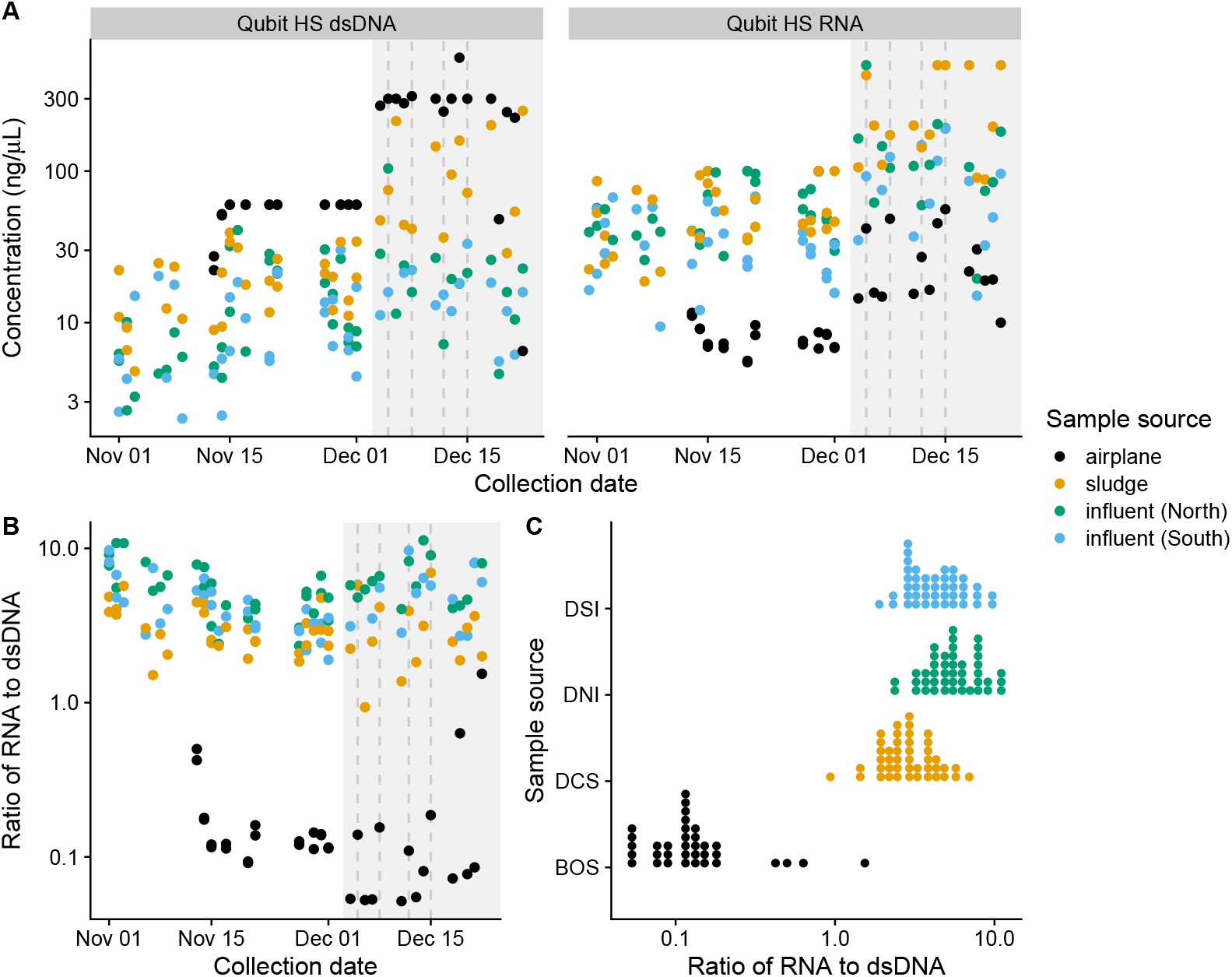
Airplane wastewater consistently yielded more double-stranded DNA (dsDNA) but less RNA than municipal wastewater. Samples collected on or after December 4 (indicated by the grey region) were measured with a different protocol which yielded higher concentration estimates; however, the differences between sample sources (Panel A, concentration in ng/µL) and the RNA:dsDNA ratios (Panel B) remained similar.

## Disclaimer

The findings and conclusions of this report are those of the authors and do not necessarily represent the official position of the U.S. Centers for Disease Control and Prevention.

## Acknowledgments

We thank Cara Seaman, the DITP sampling team, and Steve Rhode for providing samples from Deer Island Treatment Plant.

